# Structural phenotypes of osteoarthritis are clinically and genetically distinct: findings from 59,539 UK Biobank participants

**DOI:** 10.64898/2026.02.08.26345686

**Authors:** Benjamin G Faber, Mijin Jung, Raja Ebsim, Fiona R Saunders, Asad Hashmi, Sophie Scott, Jenny S Gregory, Nicholas C Harvey, John P Kemp, George Davey Smith, Andy Judge, Cindy Boer, Richard M Aspden, Claudia Lindner, Timothy Cootes, Jamie E Collins, Jonathan H Tobias

## Abstract

**OBJECTIVES:** Osteoarthritis is a heterogeneous disease, with diverse structural patterns likely reflecting distinct genetic drivers. Robust, data-driven methods to identify and characterise such phenotypes are lacking. This study leveraged the UK Biobank to define machine learning–derived structural osteoarthritis phenotypes and evaluate their clinical and genetic profiles.

**METHODS:** Machine learning models were applied to knee and hip DXA scans to derive osteophyte area, minimum joint space width, and B-scores (a combined shape vector predictive of osteoarthritis). Imaging and demographic features were clustered using k-means to classify individuals with at least one osteoarthritis feature. Phenotypes were compared with healthy controls for associations with joint pain and total joint replacement (TJR). Genetic correlations, osteoarthritis risk loci, and polygenic risk scores were analysed to define shared and distinct genetic mechanisms between phenotypes.

**RESULTS:** Among 59,539 participants (mean age 65 years; 53% female), nine reproducible phenotypes were identified, spanning joint-specific and multi-joint patterns. Hypertrophic and end-stage knee phenotypes showed the highest odds of pain (OR 7.8 [95% CI 7.1,8.7], 13.4 [9.5,19.0]) and TJR (66.0 [46.6,93.5], 127.6 [72.6,224.1]). A novel increased-cartilage phenotype was associated with greater odds of hip (3.5 [2.4,5.2]) and knee replacement (4.1 [2.6,6.6]). Distinct genetic architectures were observed; increased- and atrophic-cartilage phenotypes were inversely genetically correlated (rg −0.46 [−0.9,−0.2]) with opposing effects at *DOT1L* and *COL27A1*.

**CONCLUSIONS:** Machine learning revealed nine reproducible osteoarthritis structural phenotypes with divergent clinical and genetic signatures. These findings demonstrate that simple imaging and demographic data can stratify patients into biologically distinct phenotypes likely to require tailored treatments.

**Key messages:** **What is already known on this topic?**

- Different osteoarthritis phenotypes have been proposed, which could guide patient stratification for drug trials and pharmacotherapy. However, these proposals have mainly been based on analysis of small numbers of patients that are focused on the knee joint alone.
- To our knowledge, no systematic, hypothesis-free approach has been applied to classify different osteoarthritis phenotypes using structural features derived from large numbers of individuals.

**What this study adds?**

- This study identifies and characterises nine reproducible structural phenotypes of osteoarthritis across both the hip and knee using high-resolution DXA imaging in UK Biobank.
- It demonstrates that these phenotypes have distinct clinical profiles, with widely varying risks of joint pain and subsequent joint replacement.
- It provides robust evidence that the phenotypes differ in their genetic architecture, supporting the existence of genetically determined endotypes within osteoarthritis.

**How this study might affect research, practice or policy?**

- The findings advance understanding of the structural heterogeneity of osteoarthritis and highlight that distinct phenotypes represent different biological pathways guiding research into future disease modifying therapeutics.
- The automated, scalable methods used here could support patient stratification in clinical trials, enabling targeted evaluation of treatments in phenotypes most likely to benefit, an essential step towards a precision medicine approach in osteoarthritis.

## INTRODUCTION

Osteoarthritis is the leading cause of knee and hip pain and disability worldwide, affecting an estimated 500 million people^1^. Its prevalence continues to rise, driven by population ageing, and early osteoarthritis alone costs $106bn per year in lost economic performance and healthcare spend^2^. Despite this impact, therapeutic progress has been limited. No disease-modifying osteoarthritis drugs have yet been approved, and symptomatic treatments remain restricted to exercise therapies, analgesics and joint replacement for advanced disease^3^.

One of the major reasons for this lack of progress is the growing recognition that osteoarthritis is not a single disease but a collection of related conditions, or phenotypes, underpinned by heterogeneous biological mechanisms termed endotypes^4^. This would explain why patients with osteoarthritis can vary markedly in patterns of joint involvement, structural changes, symptom severity, and trajectories of progression^5^. This heterogeneity may dilute treatment effects in clinical trials and help to explain the failure of promising therapeutics to gain regulatory approval^6^. There is therefore a pressing need for clinically relevant phenotyping strategies that can identify subgroups of patients with shared endotypes who might respond more consistently to targeted interventions.

Multiomic analyses, including transcriptomic, proteomic, and metabolomic profiling of joint tissue, suggest that molecular endotypes exist and represent distinct biological processes within osteoarthritis^7, 8^. These endotypes provide valuable insights into disease pathways but rely on invasive tissue sampling and intensive laboratory assays that limit their scalability. Consequently, they are challenging to apply in large cohorts or in clinical settings where high-throughput stratification is required.

Parallel efforts using magnetic resonance imaging (MRI) have highlighted the existence of different structural phenotypes, such as inflammatory, atrophic, or hypertrophic patterns^5, 9^. MRI provides detailed visualisation of cartilage, bone marrow lesions, synovitis, and other features, and has contributed substantially to understanding structural variation among patients^10^. However, MRI is resource-intensive, time-consuming, and expensive to deploy at scale. Moreover, most MRI-based studies focus on a single joint, typically the knee. This limits the ability to identify broader, multi-joint phenotypes, such as systemic bone-forming patterns^11^, and constrains efforts to characterise osteoarthritis as a systemic disease^12, 13^.

UK Biobank provides a unique opportunity to address these limitations. It hosts one of the world’s largest musculoskeletal imaging initiatives, incorporating modern high-resolution dual-energy X-ray absorptiometry (DXA) of both knees and hips^14^. Although modern DXA offers less detail than MRI, its spatial resolution for bone and joint margins is good and closely resembles conventional radiography^15^. Its scalability, low cost, reduced radiation exposure, and consistent imaging protocol make it well suited for characterising structural variation across very large populations. Previous work in UK Biobank has demonstrated that radiographic osteoarthritis features derived from DXA, including joint shape measures and osteophyte formation, are predictive of future clinical outcomes such as joint pain and arthroplasty^16–18^.

Machine-learning methods have been developed to extract quantitative structural features from UK Biobank DXA images, enabling analysis at a scale not previously possible^19, 20^. These techniques quantify osteophyte area, joint space width, and joint shape in a continuous, reproducible manner, providing substantially greater resolution than traditional semi-quantitative grading^21^. In combination with UK Biobank’s extensive genetic data, this creates a powerful platform for defining structural phenotypes and linking them to underlying biological mechanisms.

The aim of the present study was to leverage this unique imaging and genetic resource to characterise structural phenotypes of osteoarthritis across both the knee and hip. By applying machine-learning–derived quantitative imaging measures and unsupervised clustering methods, we sought to identify reproducible patterns of joint structure that may reflect distinct underlying pathomechanisms, using a hypothesis-free approach. We then evaluated the clinical relevance of these phenotypes by examining associations with pain and subsequent joint replacement. Finally, we investigated their genetic architecture, testing whether phenotypes differed in genetic correlations, in the distribution of known osteoarthritis risk variants, and in polygenic risk for knee, hip, and multi-joint osteoarthritis. Through this approach, we aimed to establish an automated and high-throughput framework for scalable osteoarthritis phenotyping that could support stratification efforts in future clinical trials and ultimately contribute to the development of precision medicine in osteoarthritis.

## METHODS

### Study sample

UK Biobank is a large prospective cohort comprising 503,317 individuals aged 40–69 years at recruitment, with extensive genetic, questionnaire, clinical assessment and linked electronic health record data. Since 2014, a subset of participants has undergone high-resolution dual-energy X-ray absorptiometry (DXA) imaging of both knees and hips using a GE-Lunar iDXA scanner (Madison, WI). All participants were invited to take part in the imaging extension but exclusion criteria meant those with metal work in their body (not limbs) were excluded^14^. Ethical approval for UK Biobank was obtained from the National Information Governance Board for Health and Social Care and the North West Multi-Centre Research Ethics Committee (11/NW/0382). All participants provided written informed consent. For the present study, individuals were eligible if bilateral knee and hip DXA scans were available as of 1^st^ January 2025 (Figure 1A). Participants with evidence of partial or total joint replacement at any knee or hip joint were excluded because protheses prevent the assessment of native joint structure.

**Figure 1.**
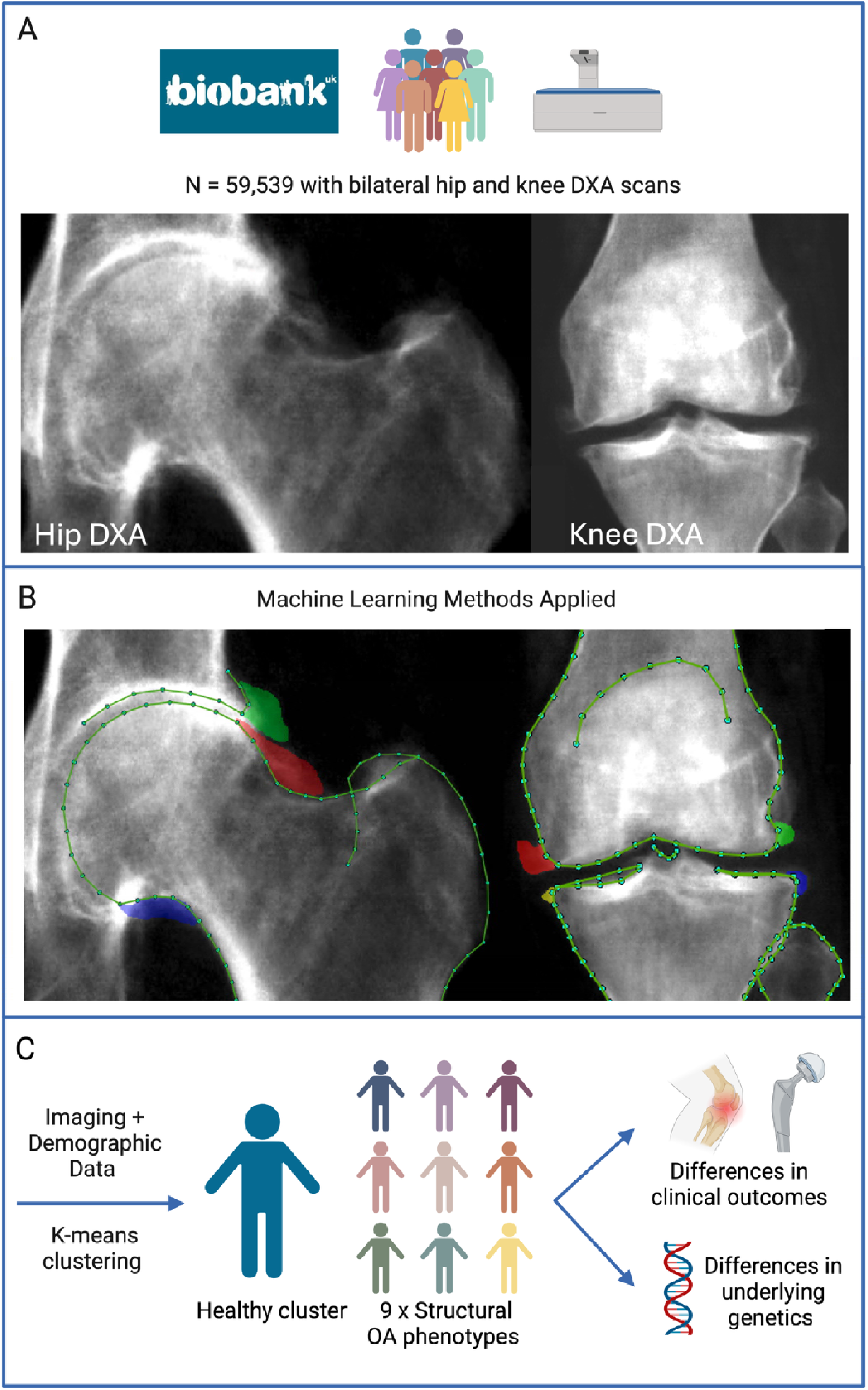
Methodological framework for study. A – example knee and hip DXA scans in UK Biobank. B – Machine learning models mark the outline of the joint with points and shade osteophytes. C – Imaging and demographic data are used to identify structural osteoarthritis (OA) phenotypes.

### Image analysis

Image analysis followed standardised protocols in which validated machine-learning algorithms were applied to each DXA image to quantify features of joint morphology and osteoarthritis (Figure 1B). These methods have been previously shown to provide accurate and reproducible measurements in the UK Biobank and Shanghai Changfeng studies^16, 17, 22^. Extracted features included osteophyte area, minimum joint space width, and the B-score, which is a shape vector that represents a composite joint shape associated with an increased risk of hospital diagnosis of osteoarthritis^18^. Measures of subchondral sclerosis and cysts could not be reliably identified on DXA and were therefore not included. All scans were manually reviewed to ensure the correct application of these methods prior to analyses.

### Phenotype identification

Structural phenotypes of osteoarthritis were derived using k-means clustering, which was selected because it is computational efficient, dealing well with continuous variables in large datasets^23^. Individuals were included in the clustering analysis if they demonstrated at least one radiographic feature of osteoarthritis at any knee or hip joint, defined as either osteophyte formation or joint space narrowing. Participants who showed no evidence of radiographic osteoarthritis at any knee or hip joint were classified as healthy controls. Prior to clustering, imaging features and demographic covariates (age, height, and weight) were standardised within sex to remove sex-driven differences in anatomical scale. The optimal number of clusters was determined using an elbow plot of within-cluster sum of squares. To assess the robustness of the clustering solution, three-fold internal validation was conducted by splitting the data into three random samples and repeating the clustering analysis on each sample. An additional sensitivity analysis repeated the clustering analysis after removing demographic variables to evaluate the influence of these characteristics on cluster formation.

### Clinical outcomes

Following cluster derivation, structural phenotypes were combined with radiographically normal controls for cross-phenotype comparisons. Clinical outcomes were evaluated using logistic regression to compare the odds of prolonged joint pain and subsequent joint replacement between each structural phenotype and the control group. Prolonged joint pain was assessed through same-day questionnaire responses to the questions “Have you had [hip/knee] pains for more than 3 months?” Subsequent total hip or knee replacement procedures were identified using linked electronic healthcare record coded with Office of Population Censuses and Survey codes (W371, W381, W391, W401, WC411, W421).

### Genetics

Genetic analyses were performed to determine whether the structural phenotypes were influenced by shared or unique genetic mechanisms. Genome-wide association studies (GWAS) were conducted separately for each phenotype using Regenie v4.1^24^, adjusting for age, sex, 20 principal components of ancestry, and genotyping chip. Analyses were restricted to individuals of European ancestry to reduce confounding from population structure. Genetic correlations between phenotypes were calculated using linkage disequilibrium score regression (LDSC)^25^ with the 1000 Genomes European reference panel. Correlations of 0.7 or higher were considered indicative of strong genetic similarity. Additional genetic correlations were estimated between each phenotype and musculoskeletal traits using GWAS data for height^26^, femoral neck bone mineral density^27^, hip cartilage thickness^28^, osteoarthritis at specific joints (i.e., knee, hip and all joints^29^), and hip shape^30^.

Due to the small nature of some clusters, discovery GWAS were underpowered. Therefore, analyses focused on 962 previously identified osteoarthritis-associated variants from the largest osteoarthritis GWAS to date^29^. Associations with each phenotype were tested, applying a 10% Benjamini–Hochberg false discovery rate across 6,516 tests. This threshold was selected given the loci had already been robustly associated with osteoarthritis and therefore the risk of false discovery risk was deemed low. Variant to gene assignments and pathway annotations were taken from Hatzikotoulas et al.^29^ and, if no functional annotation was available, the nearest gene was used.

Polygenic risk scores for knee, hip, and all-joint osteoarthritis were computed for all individuals using lead variants and effect sizes published previously^29^. Scores were generated using PLINK v1.9 and standardised to mean 0 and standard deviation 1. Differences in mean polygenic risk scores across phenotypes were analysed using analysis of variance.

### Patient and public involvement

Patient and public involvement consisted of consultation with two patient and public advisory groups, each expressing support for research into osteoarthritis stratification.

## RESULTS

### Population characteristics

A total of 59,539 UK Biobank participants met the inclusion criteria, each having bilateral knee and hip DXA imaging suitable for analysis. The mean age of the cohort was 65 years (range 45–85), the mean height was 169.7 cm (range 124–204), and the mean body weight was 75.2 kg (range 34–200) (Table 1). Females formed 52.8% of the cohort (31,450 participants). Self-reported pain was more frequently observed at the knee than at the hip, with 14.8% reporting prolonged knee pain compared with 8.3% reporting prolonged hip pain. Knee replacement was marginally more common than hip replacement (0.8% vs 0.7%), reflecting known epidemiological patterns of symptomatic and end-stage disease.

**Table 1.** Showing descriptive statistics for the study population. OP - osteophyte, mJSW - minimum joint space width, THR - total hip replacement, TKR - total knee replacement.

|  | Combined Sex<br>Mean [Range] | Female<br>Mean [Range] | Male<br>Mean [Range] |
| --- | --- | --- | --- |
| Age | 65.09 [45 - 85] | 64.49 [45 - 85] | 65.76 [46 - 85] |
| Height | 169.73 [124 - 204] | 163.38 [124 - 198] | 176.83 [140 - 204] |
| Weight | 75.20 [34 - 200] | 68.26 [34 - 200] | 82.97 [44 - 171] |
|  | Count [Percentage] | Count [Percentage] | Count [Percentage] |
| Sex (Male) | 28,089 [47.18%] | NA | NA |
|  | Mean [Range] | Mean [Range] | Mean [Range] |
| Hip B-score (Left) | -0.02 [-4.09 - 5.28] | -0.21 [-4.01 - 5.21] | 0.19 [-4.09 - 5.28] |
| Hip OP area (Left) | 2.18 [0.00 - 438.06] | 1.44 [0.00 - 367.33] | 3.02 [0.00 - 438.06] |
| Hip mJSW (Left) | 2.86 [0.02 - 5.52] | 2.79 [0.02 - 5.06] | 2.94 [0.03 - 5.52] |
| Hip B-score (Right) | 0.03 [-3.91 - 5.46] | -0.12 [-3.86 - 4.54] | 0.21 [-3.91 - 5.46] |
| Hip OP area (Right) | 2.32 [0.00 - 458.31] | 1.84 [0.00 - 312.69] | 2.85 [0.00 - 458.31] |
| Hip mJSW (Right) | 2.92 [0.01 - 6.82] | 2.83 [0.05 - 6.82] | 3.03 [0.01 - 5.24] |
| Knee B-score (Left) | 0.03 [-4.28 - 5.03] | -0.14 [-4.28 - 4.46] | 0.23 [-3.37 - 5.03] |
| Knee OP area (Left) | 3.15 [0.00 - 447.15] | 3.71 [0.00 - 447.15] | 2.53 [0.00 - 254.18] |
| Knee mJSW (Left) | 3.94 [0.01 - 7.90] | 3.64 [0.01 - 7.21] | 4.28 [0.25 - 7.90] |
| Knee B-score (Right) | -0.06 [-3.96 - 5.14] | -0.22 [-3.79 - 4.22] | 0.11 [-3.96 - 5.14] |
| Knee OP area (Right) | 2.83 [0.00 - 679.90] | 3.34 [0.00 - 447.15] | 2.25 [0.00 - 679.90] |
| Knee mJSW (Right) | 3.91 [0.19 - 8.61] | 3.63 [0.19 - 6.70] | 4.23 [0.19 - 8.61] |
|  | Count [Percentage] | Count [Percentage] | Count [Percentage] |
| Hip Pain | 4,932 [8.28%] | 3,136 [9.97%] | 1,796 [6.39%] |
| Knee Pain | 8,809 [14.80%] | 4,707 [14.97%] | 4,102 [14.60%] |
| THR | 387 [0.65%] | 228 [0.72%] | 159 [0.57%] |
| TKR | 486 [0.82%] | 262 [0.83%] | 224 [0.80%] |
| Total | 59,539 | 31,450 | 28,089 |

### Structural phenotypes

Among the 28,953 individuals who exhibited at least one radiographic feature of osteoarthritis in any knee or hip joint, k-means clustering identified nine distinct structural phenotypes (Supplementary Figure 1). Internal cluster validation revealed acceptable stability across three subsamples (Supplementary Results, Supplementary Table 1). These phenotypes were subsequently analysed alongside 30,586 radiographically normal controls (Figure 1C). Phenotype labels were assigned based on predominant structural characteristics, while recognising that each phenotype encompassed a spectrum of structural involvement (Figure 2, Supplementary Table 2).

**Figure 2.**
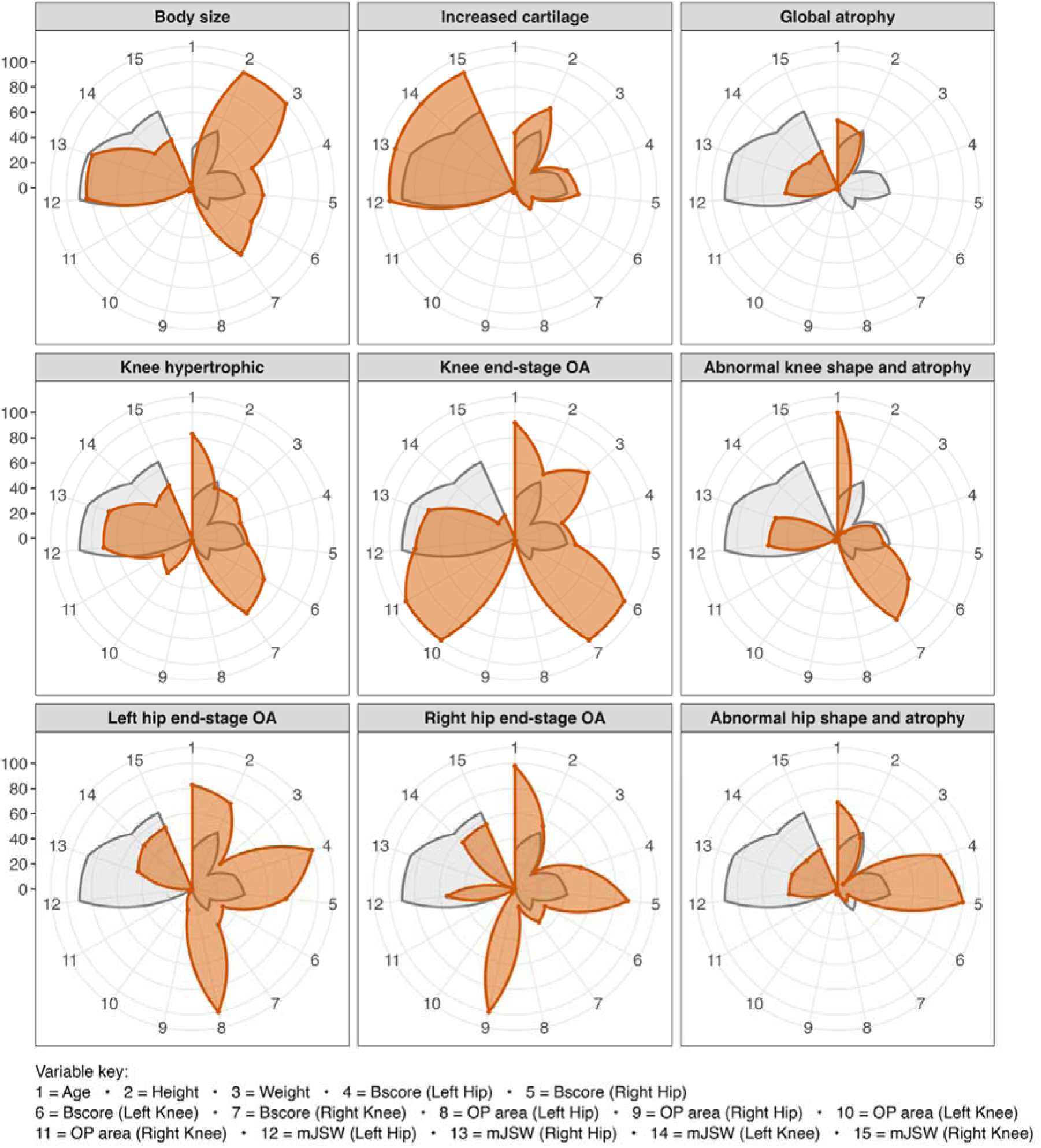
Each structural phenotype (orange) compared to healthy controls (grey) by the mean of each imaging variable (scaled 0-100) included in the k-means clustering.

Three phenotypes demonstrated multi-joint osteoarthritis patterns. The body size phenotype comprised 3,942 individuals and was characterised by above-average height and weight alongside features of osteoarthritis across multiple joints. The increased cartilage phenotype, including 5,175 individuals, was marked by relatively wide joint space in the presence of small osteophytes, suggesting a distinct pattern of thicker cartilage despite structural evidence of osteoarthritis. The global atrophy phenotype (n = 6,693) showed reduced joint space width with minimal osteophyte formation and represented a generalised atrophic pattern not specific to any single joint.

Three knee-specific phenotypes were identified. The knee hypertrophic phenotype consisted of 1,707 individuals with prominent knee osteophytes and minimal involvement at the hip. A smaller group of 134 individuals formed the knee end-stage phenotype, characterised by extensive osteophyte burden, severe joint space reduction, and clear deformity of joint shape (a higher B-score). An additional phenotype captured abnormal knee shape and cartilage atrophy, defined by high B-scores combined with narrowing of the joint space (n = 4,908).

Three hip-specific phenotypes were found. Unilateral end-stage hip osteoarthritis formed two distinct phenotypes, one for the left hip (n = 441) and one for the right (n = 418), each representing severe structural deterioration with narrowed joint space, altered joint shape and large osteophytes. A phenotype of abnormal hip shape with atrophy, defined by high B-score and joint space narrowing, was also identified (n = 5,535).

### Clinical associations

Clinical associations differed considerably across phenotypes (Table 2). Joint-specific phenotypes demonstrated the strongest associations with both pain and joint replacement. End-stage knee osteoarthritis showed 13-fold higher odds of prolonged knee pain (OR 13.4 [95% CI 9.5-19.0]) and a striking 128-fold increased odds of knee replacement compared with controls (OR 127.6 [72.6-224.1]). End-stage hip disease demonstrated similarly large associations, with odds ratios of 6.2 (95% CI 5.1-7.6) and 7.5 (6.1-9.2) for left and right hip pain, and 72.8 (50.6-104.8) and 51.7 (34.6-77.1) for left and right hip replacement, respectively. The knee hypertrophic phenotype was also strongly associated with symptoms and surgery, with odds ratios of 7.8 (7.1-8.7) for knee pain and 66.0 (46.6-93.5) for subsequent knee replacement. Shape-and-atrophy phenotypes for both knee and hip also showed substantial associations with arthroplasty, indicating the clinical importance of altered joint morphology (OR 19.6 [13.8-27.8] & 7.3 [5.3-10.1] respectively).

**Table 2.** Associations between each structural phenotype and clinical outcomes, in reference to the UK Biobank participants without signs of radiographic osteoarthritis.

| Phenotype | Hip Pain |  | Total Hip Replacement |  | Knee Pain |  | Total Knee Replacement |  |
| --- | --- | --- | --- | --- | --- | --- | --- | --- |
|  | OR [ 95% CI] | P | OR [ 95% CI] | P | OR [ 95% CI] | P | OR [ 95% CI] | P |
| Body Size | 1.42 [1.27 - 1.59] | $1.1 \times 10^{-09}$ | 2.71 [1.69 - 4.37] | $3.9 \times 10^{-05}$ | 2.66 [2.45 - 2.89] | $4 \times 10^{-121}$ | 11.05 [7.43 - 16.44] | $2 \times 10^{-32}$ |
| Increased cartilage | 1.17 [1.05 - 1.30] | $5.1 \times 10^{-03}$ | 3.51 [2.36 - 5.22] | $5.6 \times 10^{-10}$ | 1.59 [1.47 - 1.73] | $4.5 \times 10^{-28}$ | 4.10 [2.55 - 6.58] | $5.5 \times 10^{-09}$ |
| Global atrophy | 0.87 [0.78 - 0.97] | 0.01 | 0.97 [0.54 - 1.73] | 0.92 | 1.18 [1.09 - 1.28] | $5.3 \times 10^{-05}$ | 2.73 [1.66 - 4.48] | $7.3 \times 10^{-05}$ |
| Knee Hypertrophic | 1.21 [1.01 - 1.44] | 0.04 | 4.93 [2.92 - 8.32] | $2.3 \times 10^{-09}$ | 7.84 [7.08 - 8.68] | $1 \times 10^{-200}$ | 65.99 [46.59 - 93.45] | $4 \times 10^{-123}$ |
| Knee end-stage | 1.27 [0.70 - 2.30] | 0.43 | 3.48 [0.48 - 25.23] | 0.22 | 13.41 [9.45 - 19.01] | $4.9 \times 10^{-48}$ | 127.59 [72.63 - 224.11] | $7.5 \times 10^{-64}$ |
| Knee shape and atrophy | 1.28 [1.15 - 1.43] | $4.4 \times 10^{-06}$ | 3.61 [2.42 - 5.38] | $3.3 \times 10^{-10}$ | 2.73 [2.53 - 2.94] | $2 \times 10^{-152}$ | 19.63 [13.84 - 27.84] | $1.5 \times 10^{-62}$ |
| Left hip end-stage | 6.19 [5.05 - 7.60] | $1.1 \times 10^{-68}$ | 72.82 [50.61 - 104.79] | $5 \times 10^{-118}$ | 2.00 [1.57 - 2.54] | $1.6 \times 10^{-08}$ | 11.73 [5.24 - 26.26] | $2.1 \times 10^{-09}$ |
| Right hip end-stage | 7.53 [6.14 - 9.23] | $2.7 \times 10^{-84}$ | 51.65 [34.62 - 77.06] | $3.2 \times 10^{-83}$ | 1.77 [1.37 - 2.29] | $1.3 \times 10^{-05}$ | 8.80 [3.47 - 22.37] | $4.8 \times 10^{-06}$ |
| Hip shape and atrophy | 1.55 [1.41 - 1.70] | $4.3 \times 10^{-19}$ | 7.30 [5.29 - 10.07] | $1.1 \times 10^{-33}$ | 1.37 [1.26 - 1.49] | $3.9 \times 10^{-13}$ | 3.70 [2.29 - 5.97] | $8.8 \times 10^{-08}$ |

Multi-joint phenotypes showed more moderate yet clinically relevant associations. The body-size phenotype displayed increased risk of both knee (OR 2.7 [2.5-2.9]) and hip pain (OR 1.4 [1.3-1.6]), and a notably higher risk of knee replacement (OR 11.1 [7.4-16.4]) than hip replacement (OR 2.7 [1.7-4.4]). The increased cartilage phenotype demonstrated modest but consistently elevated risk of both symptoms (Hip OR 1.2 [1.1-1.3] & Knee OR 1.6 [1.5-1.7]) and arthroplasty (Hip OR 3.5 [2.4-5.2] & Knee OR 4.14 [2.6-6.6]) across both joints. The global atrophy phenotype exhibited little association with hip outcomes (Pain OR 0.9 [0.8-1.0] & joint replacement OR 1.0 [0.5-1.7]) but a small increase in risk of knee pain (OR 1.2 [1.1-1.3]) and replacement (OR 2.7 [1.7-4.5]).

### Genetic correlations

Genetic correlations between phenotypes revealed divergent genetic architectures (Figure 3; Supplementary Table 3). No pair of phenotypes exhibited high genetic correlation (rg ≥ 0.7), suggesting the potential presence of unique genetic contributions for each structural phenotype of osteoarthritis alongside shared factors. Increased cartilage and global atrophy showed a negative correlation (rg −0.46 [95% CI −0.9,−0.2]), consistent with their opposing phenotypic features. The strongest positive correlation was between increased cartilage and knee hypertrophic osteoarthritis (rg 0.67 [0.1,1.2]), suggesting shared anabolic biological pathways.

**Figure 3.**
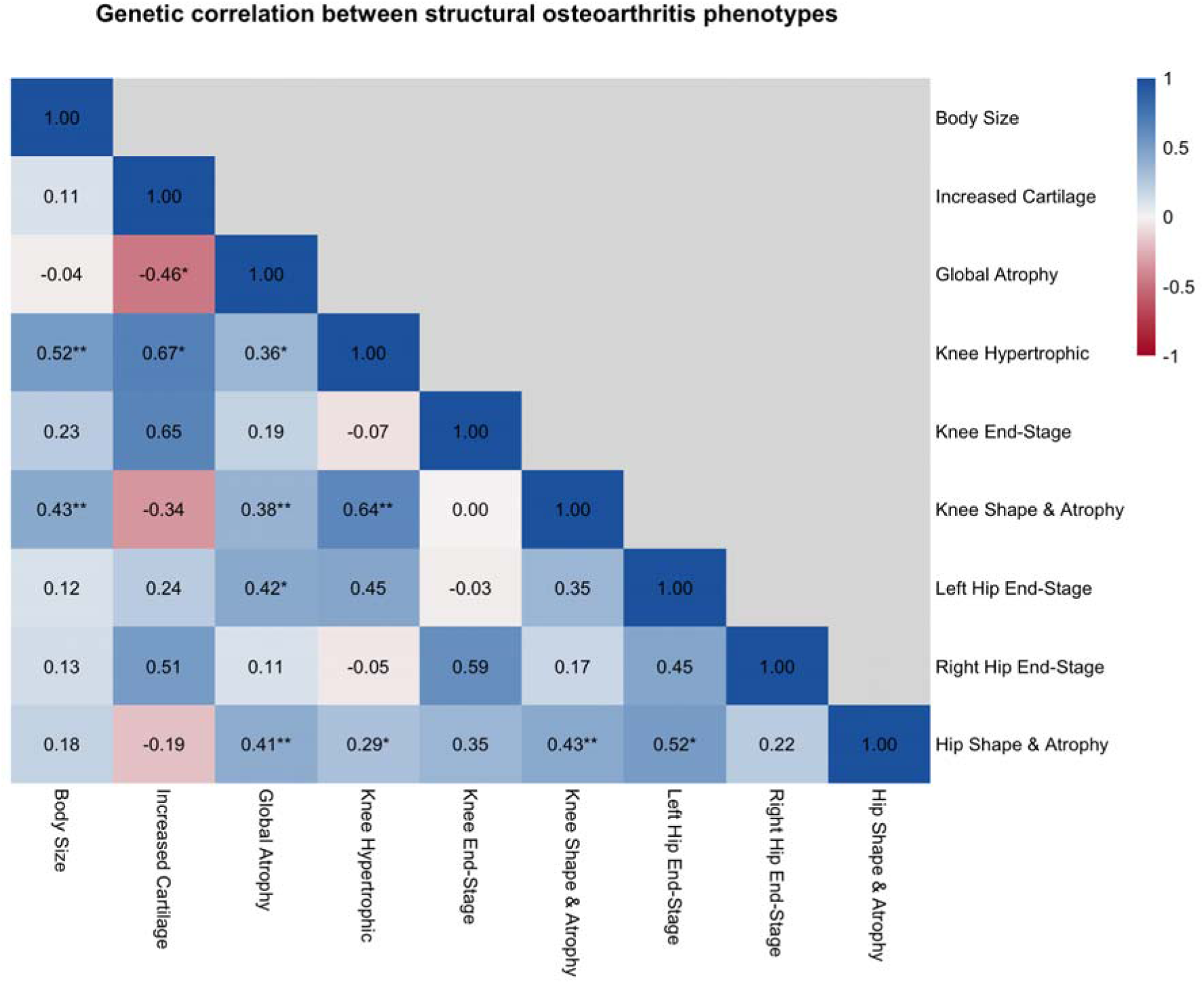
A correlation matrix showing the genetic correlation estimates between structural phenotypes. * p-value <0.05, ** p-value <0.001

Genetic correlations between structural phenotypes and musculoskeletal traits are shown in Supplementary Table 4. Knee and hip based structural phenotypes derived here from imaging were most strongly correlated with clinically defined osteoarthritis at the same joint. Both the body-size and increased-cartilage phenotypes showed strong positive genetic correlations with height (rg 0.45 [0.4,0.5] & 0.47 [0.2,0.8] respectively). The knee hypertrophic phenotype was uniquely correlated with bone mineral density (rg 0.24 [0.03,0.4]), consistent with the hypothesis that osteophyte formation is linked with a global tendency towards bone formation. Global atrophy demonstrated a strong negative correlation with hip cartilage thickness (rg −0.77 [−0.9,−0.6]).

### Genetic associations

Of the 962 known osteoarthritis variants examined, 92 showed evidence of associations (Benjamini-Hochberg adjusted P<0.1) with at least one structural phenotype (Supplementary Table 5). Phenotypes with larger numbers, such as body size and increased cartilage, had more associated variants, whereas relatively few individuals had end-stage phenotypes, limiting statistical power. Fine mapping suggested that several genes were associated with multiple phenotypes (Figure 4). Genes implicated in increased cartilage and the atrophic phenotypes, *DOT1L, COL27A1, C2orf40* and *IL11,* reassuringly showed opposing directions of effect. In addition, pathway-specific genes were identified, relating to fibroblast growth factor (FGF), transforming growth factor beta (TGF-β), wingless-related integration site (Wnt), extracellular matrix (ECM), and bone morphogenetic protein (BMP) pathways (Figure 4).

**Figure 4.**
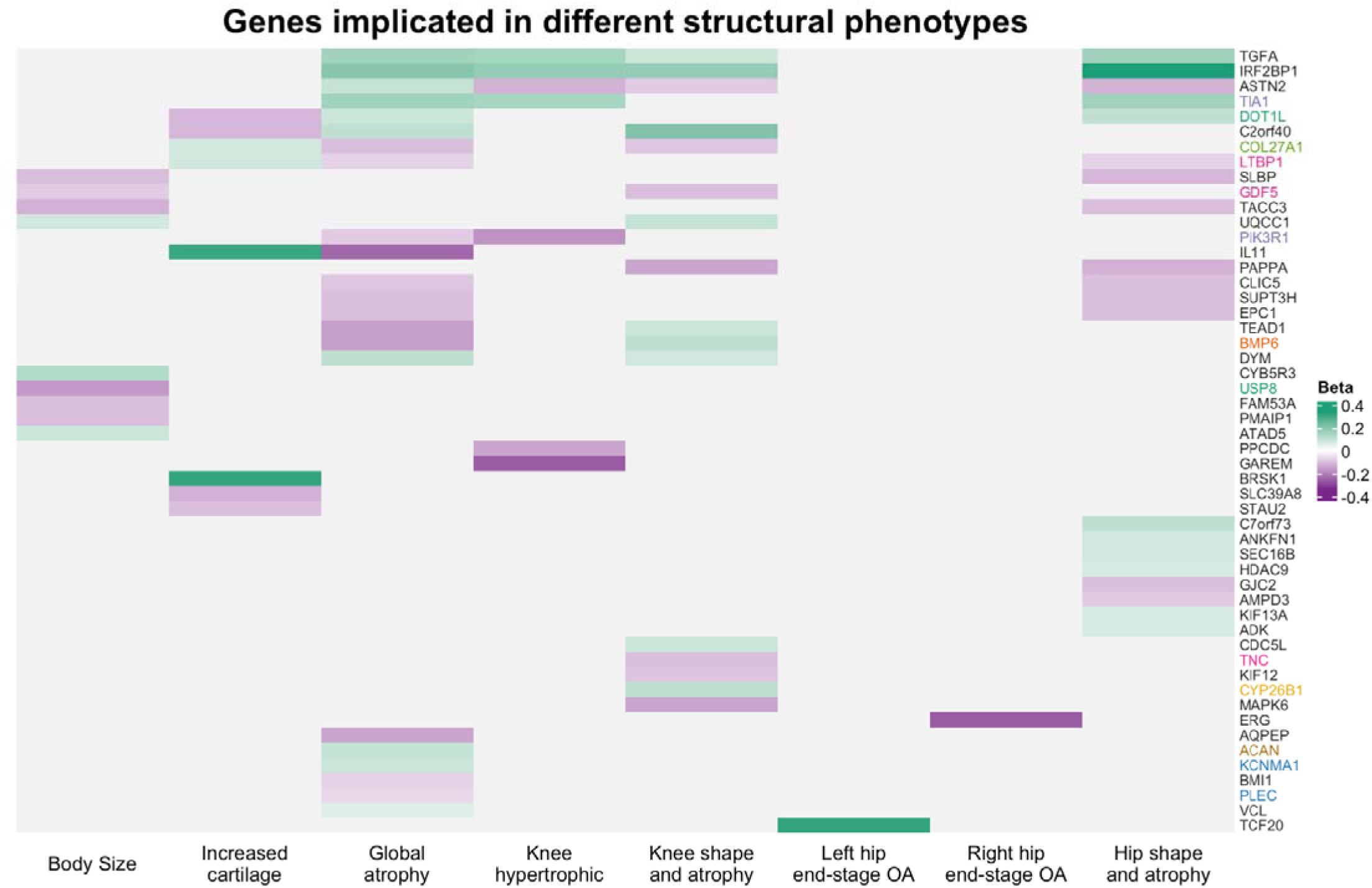
Osteoarthritis genes implicated in each structural phenotype through variant look up. Horizontal bars represent variant betas, which were used to compare effects between phenotypes (the colour represents the effect size as indicated by the legend). Gene names are coloured according to common pathways: Royal blue – Circadian Rhythm, Teal green - Wingless-related integration site (Wnt), Pink-magenta – Transforming Growth Factor Beta (TGFB), Violet – Fibroblast Growth Factor (FGF), Leaf green – Extracellular matrix (ECM), Orange – Bone morphogenetic protein (BMP), Yellow – Retinoic Acid, Brown – FGF & ECM

### Polygenic risk scores

Polygenic risk scores further supported the distinction between different phenotypes (Table 3). The highest knee polygenic risk scores were observed in knee hypertrophic (mean 0.19 SD) and knee end-stage (0.16) phenotypes, with no corresponding elevation in hip score. Conversely, left and right end-stage hip phenotypes showed the highest hip polygenic risk scores (0.23 & 0.29 respectively). Most other phenotypes displayed modest elevations in polygenic risk scores, except mild atrophy, which demonstrated reduced polygenic risk across traits.

**Table 3.** Polygenic risk score (PRS) for hip, knee and multi joint osteoarthritis (OA). Bold values are greater than 0.1 standard deviation difference from the mean. “All OA” means all joint definitions of osteoarthritis were included in the underlying genome-wide association study. ^29^.

|  | Hip OA PRS | Knee OA PRS | All OA PRS |
| --- | --- | --- | --- |
| Phenotype | Mean PRS [Range] | Mean PRS [Range] | Mean PRS [Range] |
| Healthy | -0.02 [ -4.34 , 4.29] | -0.03 [ -4.39 , 4.05] | -0.04 [ -4.42 , 4.62] |
| Body Size | 0.03 [ -3.19 , 3.52] | 0.07 [ -3.48 , 3.18] | 0.05 [ -3.22 , 3.80] |
| Increased Cartilage | 0.04 [ -3.11 , 4.33] | 0.02 [ -3.61 , 3.16] | 0.01 [ -3.56 , 3.54] |
| Global Atrophy | -0.07 [ -3.48 , 3.97] | -0.04 [ -3.70 , 3.24] | -0.05 [ -4.58 , 3.48] |
| Knee Hypertrophic | 0.03 [ -3.38 , 3.95] | **0.19 [ -2.97 , 4.59]** | **0.11 [ -3.02 , 3.55]** |
| Knee End-stage | -0.09 [ -2.59 , 2.36] | **0.16 [ -2.48 , 2.36]** | 0.00 [ -2.76 , 2.70] |
| Knee Shape and Atrophy | -0.02 [ -3.24 , 3.75] | 0.03 [ -3.37 , 3.73] | 0.05 [ -4.01 , 3.26] |
| Left Hip End-stage | **0.23 [ -2.86 , 3.32]** | 0.04 [ -2.80 , 3.21] | **0.21 [ -2.62 , 3.19]** |
| Right Hip End-stage | **0.29 [ -2.72 , 3.22]** | 0.05 [ -3.18 , 2.89] | **0.23 [ -3.46 , 2.87]** |
| Hip Shape and Atrophy | 0.01 [ -3.84 , 3.52] | -0.06 [ -4.03 , 3.79] | 0.02 [ -3.64 , 3.75] |
| P-value from ANOVA | $1.41 \times 10^{-20}$ | $1.29 \times 10^{-25}$ | $5.03 \times 10^{-27}$ |

Figure 5 summarises the clinical and genetic profiles of all structural phenotypes.

**Figure 5.**
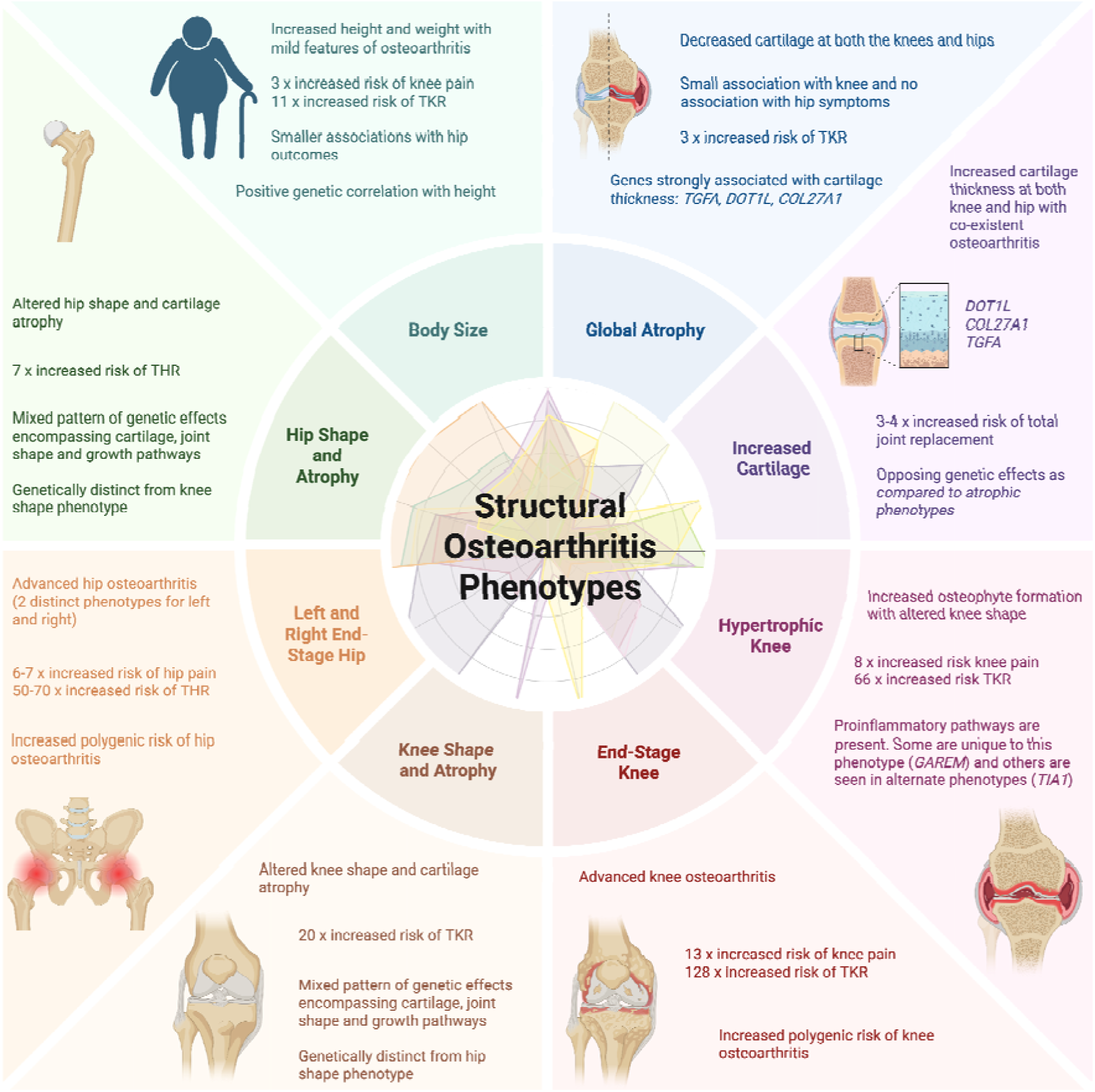
A summary overview of the structural phenotypes of osteoarthritis. Their construct, clinical and genetic profiles. Left and right end-stage hip osteoarthritis are combined for display purposes. (THR = Total Hip Replacement, TKR = Total Knee Replacement).

## DISCUSSION

This study leveraged the unprecedented scale of UK Biobank’s imaging resources to identify nine structural phenotypes of knee and hip osteoarthritis, each defined by quantitative imaging features and demographics, and distinguished by unique clinical and genetic characteristics. These findings provide strong support for the argument that osteoarthritis is not a single disease but a collection of biologically distinct structural phenotypes^4^. The ability to identify these phenotypes using scalable, automated methods applied to simple two-dimensional imaging highlights a feasible route towards incorporating structural phenotyping into future clinical care.

Traditional radiographic assessment of osteoarthritis relies heavily on semi-quantitative scoring systems that grade osteophytes and joint space narrowing as discrete categories^21^. Although well established, these methods reduce complex structural variation to simplified scales that can obscure important differences between individuals^10, 31^. By extracting and subsequently applying continuous, high-resolution measures of osteophyte area, minimum joint space width, and joint shape via B-scores, this study was able to capture subtle but clinically meaningful differences in joint structures^16–18^. This approach aligns with the recognition that osteoarthritis features exist along a continuum, and that quantitative measures better reflect disease mechanisms^32^.

The clustering analysis identified phenotypes that align with previously hypothesised forms of osteoarthritis, such as hypertrophic and metabolic disease, while also revealing unexpected phenotypes, including the increased cartilage phenotype^5^. The knee hypertrophic phenotype displayed prominent osteophyte formation with maintained joint space, and strong associations with pain and joint replacement, supporting long-standing observations that hypertrophic remodelling reflects a biologically active form of disease^33^. Its strong clinical associations and distinct genetic profile, including genetic correlation with bone mineral density and enrichment for FGF-pathway genes, reinforce the concept that osteophytosis is, at least partly, genetically determined^11, 34^. The presence of genes such as *GAREM* and *TIA1*, which are implicated in synovial inflammation^35, 36^, suggests that synovitis may be particularly relevant to this phenotype.

The body size phenotype resembles the previously described metabolic phenotype, in which higher body mass is accompanied by low-grade systemic inflammation and increased susceptibility to symptoms^37^. Although this study did not assess measures of inflammation, it is notable that the body size phenotype, characterised by substantially greater average weight, showed higher odds of knee pain and knee replacement than of hip outcomes. This pattern is consistent with established evidence that excess body weight imposes a greater mechanical and pathological burden on the knee than on the hip^38^.

Other notable structural phenotypes included the identification of end-stage osteoarthritis subtypes at both the knee and hip. These phenotypes exhibited the largest odds ratios for pain and joint replacement, underscoring the clinical significance of advanced structural deterioration^39^. The high joint-specific polygenic risk scores observed in these phenotypes suggests that genetic predisposition influences not only the occurrence of osteoarthritis but also the likelihood of progressing to severe disease. This observation provides a key link between genetic risk and the structural trajectory of osteoarthritis, supporting a model in which genetically determined biological pathways influence structural deterioration^40^.

An unexpected finding was the increased cartilage phenotype, defined by wider joint space width despite the presence of osteophytes. Joint space loss is widely considered a hallmark radiographic feature of osteoarthritis, yet this opposing phenotype demonstrated a three to four-fold increased risk of both knee and hip replacement. Previous studies found joint space width was not a good predictor of symptoms, which might be explained by the presence of this previously unrecognised opposing phenotype^41, 42^. Observed opposing genetic associations between the increased cartilage and atrophic phenotypes for *DOT1L* and *COL27A1*, which have all been previously implicated in joint space width^28^, reinforce the biological plausibility of this phenotype. Its strong positive genetic correlation with height further suggests involvement of systemic growth pathways. The pathomechanism of this phenotype might mirror acromegaly, where excess growth hormone leads to widened joint space and early osteoarthritis due to altered biomechanics and joint instability^43^. Longitudinal studies are needed to understand how this phenotype progresses.

Altered joint shape emerged as another important structural characteristic. B-scores quantify deviations from healthy joint morphology and have been shown to predict progression in knee osteoarthritis when derived from both MRI and DXA scans^18, 31^. In this study, phenotypes characterised primarily by abnormal shape and cartilage atrophy at either the knee or hip were consistently associated with increased risk of pain and arthroplasty. Their genetic associations included *GDF5*, a key regulator of joint development known to influence both osteoarthritis and height^44, 45^. This highlights how developmental pathways overlap with susceptibility to osteoarthritis and supports the hypothesis that joint morphology is a major component of disease progression^46, 47^.

The genetic analyses provide compelling evidence that the structural phenotypes develop as a result of biologically distinct subgroups (endotypes). No pair of phenotypes exhibited high genetic correlation, and polygenic risk score profiles differed across nearly all phenotypes. These findings parallel multiomic studies showing that pathways related to inflammation, cartilage remodelling, bone turnover, and growth are variably expressed among individuals with osteoarthritis^7, 8^. Whereas earlier studies often focused on identifying molecular endotypes using tissue samples or blood biomarkers, the current study demonstrates that genetically distinct endotypes can be identified directly from imaging, without the need for invasive sampling. This bridge between structural phenotype and genetically driven endotype is a key advance, illustrating that routine imaging contains information reflective of underlying biological processes.

The implications for clinical research and therapeutic development are substantial. The ability to accurately phenotype patients could help overcome the obstacle of osteoarthritis heterogeneity in trials of disease-modifying drugs. Stratifying individuals according to structural phenotype may allow trials to target specific biological mechanisms in patient groups most likely to benefit, increasing the probability of demonstrating efficacy^4, 6^. For example, sprifermin, a recombinant FGF18 analogue aimed at stimulating cartilage formation, might be more effective in phenotypes associated with FGF-pathway enrichment, such as shape-and-atrophy or global atrophy phenotypes^48^. Conversely, treatments that stimulate cartilage growth may be unsuitable for individuals with increased cartilage phenotypes.

The study’s population-based design ensures broad representation but also integrates both disease variation and disease stage into the clustering model. Although some phenotypes, particularly the end-stage groups, clearly captured advanced disease, the distinct genetic profiles observed across phenotypes suggest that differences in underlying biology, rather than merely disease severity or duration, are the primary drivers. The inclusion of both hip and knee imaging within the same analytical framework provided a comprehensive perspective not achievable in studies focused on a single joint^7, 9^. The finding that both joint specific and multi-joint osteoarthritis phenotypes exist could have relevance for clinical care by suggesting instances for screening other joints for osteoarthritis.

Several limitations must be acknowledged. First, DXA does not capture inflammatory features such as synovitis, which may play a key role in symptomatology and treatment response. Although some structural features, such as those seen in the hypertrophic phenotype, may indirectly reflect inflammatory pathways, further work is needed to determine how best to identify patients who would benefit from MRI evaluation. Second, the absence of external validation limits the generalisability of these findings. While internal validation was reassuring, some phenotypes that included fewer cases were less stable, underscoring the importance of large datasets for capturing the full heterogeneity of osteoarthritis. Thirdly, k-means requires the number of clusters to be prespecified, which can be difficult to define. That said, it is transparent partitioning method that provides a computationally efficient, stable, and parsimonious approach for large sample sizes. Finally, this study did not evaluate longitudinal data other than joint replacement; incorporating detailed follow-up imaging, which will shortly become possible as UK Biobank plans to rescan ∼60,000 participants, would allow a more nuanced understanding of structural progression within phenotypes. Previous work has suggested molecular endotypes of knee osteoarthritis are longitudinally stable but it is likely patients will transition from the structural phenotypes to end-stage disease at different rates^49^.

In summary, this study identified nine structural phenotypes of osteoarthritis with distinct clinical and genetic signatures using scalable quantitative analysis of routine DXA imaging. These phenotypes capture meaningful biological variation, have differing associations with pain and surgical outcomes, and reflect genetic pathways that are increasingly becoming therapeutic targets. The findings demonstrate that structural phenotyping offers a viable approach to stratifying patients and a promising route toward personalised care in osteoarthritis. Future work should focus on determining how treatment response varies across these phenotypes and how imaging-based stratification can be integrated into trial design and clinical decision-making.

## Data Availability

All UK Biobank data (project number 17295) will be available via their data showcase shortly after publication.

## Acknowledgements and affiliations

The authors would like to acknowledge the huge contribution of Dr Monika Frysz and Dr Rhona Beynon in establishing this unprecedented imaging resource in UK Biobank. Both individuals declined the offer of authorship as they have left academia. We would like to thank all UK Biobank participants and the members of My Friday Coffee Morning and PEP-R groups at the University of Bristol who provided input on this work. Figures were created in https://BioRender.com.

## Declaration of generative AI and AI-assisted technologies in the manuscript preparation process

During the preparation of this work BGF used Chat-GPT 5 in order to refine code to prepare the figures, complete the analyses and edit text. After using this tool, BGF reviewed and edited all of the output and takes full responsibility for the content of the published article.

## Author contributions

Authors were involved in the following elements of the study:

- Conceptualization: BGF, AJ, JK, GDS, CB, JC, JHT
- Data curation: BGF MJ RE FRS AH SS
- Formal analysis: BGF MJ RE FRS AH
- Funding acquisition: BGF JG NH JK GDS AJ CB RA CL TC JC JHT
- Investigation: BGF MJ RE FRS AH SS JC
- Methodology: BGF RE FRS JG NH JK GDS AJ CB RA CL TC JC JHT
- Software: RE CL TC
- Supervision: NH JK GDS AJ CB RA CL TC JC JHT
- Validation: BGF MJ RE FRS
- Visualization: BGF MJ RE FRS AH SS
- Writing – original draft: BGF JHT
- Writing – review and editing: BGF MJ RE FRS AH SS JG NH JK GDS AJ CB RA CL TC JC JHT

## References

1. Disease GBD, Injury, Risk Factor C. Burden of 375 diseases and injuries, risk-attributable burden of 88 risk factors, and healthy life expectancy in 204 countries and territories, including 660 subnational locations, 1990-2023: a systematic analysis for the Global Burden of Disease Study 2023. Lancet. 2025;406(10513):1873–922.

2. Weng Q, Chen Q, Jiang T, Zhang Y, Zhang W, Doherty M, et al. Global burden of early-onset osteoarthritis, 1990–2019: results from the Global Burden of Disease Study 2019. Annals of the Rheumatic Diseases. 2024;83(7):915.

3. Hunter DJ, Bierma-Zeinstra S. Osteoarthritis. Lancet. 2019;393(10182):1745–59.

4. Hunter DJ, Deveza LA. Deconstructing the “types” of osteoarthritis. Osteoarthritis Imaging. 2025;5(1):100257.

5. Roemer FW, Jarraya M, Collins JE, Kwoh CK, Hayashi D, Hunter DJ, et al. Structural phenotypes of knee osteoarthritis: potential clinical and research relevance. Skeletal Radiol. 2023;52(11):2021–30.

6. Li S, Cao P, Chen T, Ding C. Latest insights in disease-modifying osteoarthritis drugs development. Ther Adv Musculoskelet Dis. 2023;15:1759720X231169839.

7. Rockel JS, Sharma D, Espin-Garcia O, Hueniken K, Sandhu A, Pastrello C, et al. Deep learning–based clustering for endotyping and post-arthroplasty response classification using knee osteoarthritis multiomic data. Annals of the Rheumatic Diseases. 2025;84(5):844–55.

8. Angelini F, Widera P, Mobasheri A, Blair J, Struglics A, Uebelhoer M, et al. Osteoarthritis endotype discovery via clustering of biochemical marker data. Ann Rheum Dis. 2022;81(5):666–75.

9. Liu Y, Xing Z, Wu B, Chen N, Wu T, Cai Z, et al. Association of MRI-based knee osteoarthritis structural phenotypes with short-term structural progression and subsequent total knee replacement. Journal of Orthopaedic Surgery and Research. 2024;19(1):699.

10. Roemer FW, Guermazi A, Demehri S, Wirth W, Kijowski R. Imaging in Osteoarthritis. Osteoarthritis and Cartilage. 2022;30(7):913–34.

11. Hartley A, Hardcastle SA, Paternoster L, McCloskey E, Poole KES, Javaid MK, et al. Individuals with high bone mass have increased progression of radiographic and clinical features of knee osteoarthritis. Osteoarthritis Cartilage. 2020;28(9):1180–90.

12. Nelson AE. Multiple joint osteoarthritis (MJOA): What’s in a name? Osteoarthritis Cartilage. 2024;32(3):234–40.

13. Collins KH, Haugen IK, Neogi T, Guilak F. Osteoarthritis as a systemic disease. Nature Reviews Rheumatology. 2026;22(2):105–17.

14. Littlejohns TJ, Holliday J, Gibson LM, Garratt S, Oesingmann N, Alfaro-Almagro F, et al. The UK Biobank imaging enhancement of 100,000 participants:[rationale, data collection, management and future directions. Nature Communications. 2020;11(1):2624.

15. Yoshida K, Barr RJ, Galea-Soler S, Aspden RM, Reid DM, Gregory JS. Reproducibility and Diagnostic Accuracy of Kellgren-Lawrence Grading for Osteoarthritis Using Radiographs and Dual-Energy X-ray Absorptiometry Images. J Clin Densitom. 2015;18(2):239–44.

16. Beynon RA, Saunders FR, Ebsim R, Faber BG, Jung M, Gregory JS, et al. A novel classifier of radiographic knee osteoarthritis for use on knee DXA images is predictive of joint replacement in UK Biobank. Rheumatol Adv Pract. 2025;9(1):rkaf009.

17. Faber BG, Ebsim R, Saunders FR, Frysz M, Lindner C, Gregory JS, et al. A novel semi-automated classifier of hip osteoarthritis on DXA images shows expected relationships with clinical outcomes in UK Biobank. Rheumatology (Oxford). 2021;Aug 30;61(9):3586–3595.

18. Beynon RA, Saunders FR, Ebsim R, Frysz M, Faber BG, Gregory JS, et al. Dual-energy X-ray absorptiometry derived knee shape may provide a useful imaging biomarker for predicting total knee replacement: Findings from a study of 37,843 people in UK Biobank. Osteoarthr Cartil Open. 2024;6(2):100468.

19. Frysz M, Faber BG, Ebsim R, Saunders FR, Lindner C, Gregory JS, et al. Machine-learning derived acetabular dysplasia and cam morphology are features of severe hip osteoarthritis: findings from UK Biobank. J Bone Miner Res. 2022;Sep;37(9):1720–1732.

20. Ebsim R, Faber BG, Saunders F, Frysz M, Gregory J, Harvey NC, et al., editors. Automatic Segmentation of Hip Osteophytes in DXA Scans Using U-Nets. Medical Image Computing and Computer Assisted Intervention – MICCAI 2022; 2022 2022//; Cham: Springer Nature Switzerland.

21. Kellgren JH, Lawrence JS. Radiological assessment of osteo-arthrosis. Ann Rheum Dis. 1957;16(4):494–502.

22. Zheng J, Frysz M, Faber BG, Lin H, Ebsim R, Ge J, et al. Comparison between UK Biobank and Shanghai Changfeng suggests distinct hip morphology may contribute to ethnic differences in the prevalence of hip osteoarthritis. Osteoarthritis Cartilage. 2023;5:S1063-4584(23)00958-5.

23. Chin S, Collins JE. Clustering Methods in Rheumatic and Musculoskeletal Disease Research: An Educational Guide to Best Research Practices. J Rheumatol. 2024;51(12):1160–8.

24. Mbatchou J, Barnard L, Backman J, Marcketta A, Kosmicki JA, Ziyatdinov A, et al. Computationally efficient whole-genome regression for quantitative and binary traits. Nature Genetics. 2021;53(7):1097–103.

25. Bulik-Sullivan B, Finucane HK, Anttila V, Gusev A, Day FR, Loh PR, et al. An atlas of genetic correlations across human diseases and traits. Nat Genet. 2015;47(11):1236–41.

26. Yengo L, Vedantam S, Marouli E, Sidorenko J, Bartell E, Sakaue S, et al. A saturated map of common genetic variants associated with human height. Nature. 2022;610(7933):704–12.

27. Zheng HF, Forgetta V, Hsu YH, Estrada K, Rosello-Diez A, Leo PJ, et al. Whole-genome sequencing identifies EN1 as a determinant of bone density and fracture. Nature. 2015;526(7571):112–7.

28. Faber BG, Frysz M, Boer CG, Evans D, Ebsim R, Flynn K, et al. The identification of distinct protective and susceptibility mechanisms for hip osteoarthritis: findings from a genome-wide association study meta-analysis of minimum joint space width and Mendelian randomisation cluster analyses. eBioMedicine 2023;Sep;95:104759.

29. Hatzikotoulas K, Southam L, Stefansdottir L, Boer CG, McDonald ML, Pett JP, et al. Translational genomics of osteoarthritis in 1,962,069 individuals. Nature. 2025.

30. Faber BG, Frysz M, Zheng J, Lin H, Flynn KA, Ebsim R, et al. The genetic architecture of hip shape and its role in the development of hip osteoarthritis and fracture. Human Molecular Genetics. 2024;Nov 22:ddae169.

31. Bowes MA, Kacena K, Alabas OA, Brett AD, Dube B, Bodick N, et al. Machine-learning, MRI bone shape and important clinical outcomes in osteoarthritis: data from the Osteoarthritis Initiative. Ann Rheum Dis. 2021;80(4):502–8.

32. Teoh YX, Lai KW, Usman J, Goh SL, Mohafez H, Hasikin K, et al. Discovering Knee Osteoarthritis Imaging Features for Diagnosis and Prognosis: Review of Manual Imaging Grading and Machine Learning Approaches. J Healthc Eng. 2022;2022:4138666.

33. van der Kraan PM, van den Berg WB. Osteophytes: relevance and biology. Osteoarthritis and Cartilage. 2007;15(3):237–44.

34. Zhou S, Wang Z, Tang J, Li W, Huang J, Xu W, et al. Exogenous fibroblast growth factor 9 attenuates cartilage degradation and aggravates osteophyte formation in post-traumatic osteoarthritis. Osteoarthritis Cartilage. 2016;24(12):2181–92.

35. Wu Z, Liu Q, Cao Z, Li H, Zhou Y, Zhang P. Icariin decreases cell proliferation and inflammation of rheumatoid arthritis-fibroblast like synoviocytes via GAREM1/MAPK signaling pathway. Immunopharmacol Immunotoxicol. 2024;46(1):86–92.

36. Phillips K, Kedersha N, Shen L, Blackshear PJ, Anderson P. Arthritis suppressor genes TIA-1 and TTP dampen the expression of tumor necrosis factor alpha, cyclooxygenase 2, and inflammatory arthritis. Proc Natl Acad Sci U S A. 2004;101(7):2011–6.

37. Calvet J, Garcia-Manrique M, Berenguer-Llergo A, Orellana C, Cirera SG, Llop M, et al. Metabolic and inflammatory profiles define phenotypes with clinical relevance in female knee osteoarthritis patients with joint effusion. Rheumatology (Oxford). 2023;62(12):3875–85.

38. Hussain SM, Wang Y, Shaw JE, Wluka AE, Graves S, Gambhir M, et al. Relationship of weight and obesity with the risk of knee and hip arthroplasty for osteoarthritis across different levels of physical performance: a prospective cohort study. Scandinavian Journal of Rheumatology. 2019;48(1):64–71.

39. Poole KES, Burkov IS, Treece GM, Gee AH, Johannesdottir F, D’Amore S, et al. The clinical utility of imaging in osteoarthritis and its importance in future prediction of total hip replacement; a nested case-control study within the AGES-Reykjavik cohort. J Bone Miner Res. 2025.

40. Boer CG. Osteoarthritis year in review 2024: Genetics, genomics, and epigenetics. Osteoarthritis Cartilage. 2025;33(1):50–7.

41. Lanyon P, O’Reilly S, Jones A, Doherty M. Radiographic assessment of symptomatic knee osteoarthritis in the community: definitions and normal joint space. Ann Rheum Dis. 1998;57(10):595–601.

42. Faber BG, Ebsim R, Saunders FR, Frysz M, Lindner C, Gregory JS, et al. Osteophyte size and location on hip DXA scans are associated with hip pain: findings from a cross sectional study in UK Biobank. Bone. 2021:116146.

43. Poudel SB, Ruff RR, Yildirim G, Dixit M, Michot B, Gibbs JL, et al. Excess Growth Hormone Triggers Inflammation-Associated Arthropathy, Subchondral Bone Loss, and Arthralgia. Am J Pathol. 2023;193(6):829–42.

44. Coveney CR, Maridas D, Chen H, Muthuirulan P, Liu Z, Jagoda E, et al. Complex Regulatory Interactions at GDF5 Shape Joint Morphology and Osteoarthritis Disease Risk. Arthritis Rheumatol. 2025;77(11):1488–502.

45. Sanna S, Jackson AU, Nagaraja R, Willer CJ, Chen WM, Bonnycastle LL, et al. Common variants in the GDF5-UQCC region are associated with variation in human height. Nat Genet. 2008;40(2):198–203.

46. Croft P, Cooper C, Wickham C, Coggon D. Defining osteoarthritis of the hip for epidemiologic studies. American Journal of Epidemiology. 1990;132(3):514–22.

47. Faber BG, Frysz M, Hartley AE, Ebsim R, Boer CG, Saunders FR, et al. A Genome-Wide Association Study Meta-Analysis of Alpha Angle Suggests Cam-Type Morphology May Be a Specific Feature of Hip Osteoarthritis in Older Adults. Arthritis Rheumatol. 2023;75(6):900–9.

48. Bay-Jensen AC, Manginelli A, Karsdal MA, Luo Y, He Y, Michaelis M, et al. A low repair endotype is predictive for response to FGF-18 treatment: data from the phase II oa forward study. Osteoarthritis and Cartilage. 2021;29:S144.

49. Hannani MT, Thudium CS, Gellhorn AC, Larkin J, Karsdal MA, Lisowska-Petersen Z, et al. Longitudinal stability of molecular endotypes of knee osteoarthritis patients. Osteoarthritis Cartilage. 2025;33(1):166–75.

