## Supplementary for "Structural phenotypes of osteoarthritis are clinically and genetically distinct: findings from 59,539 UK Biobank participants": Supplementary Figures 17.11.25.docx

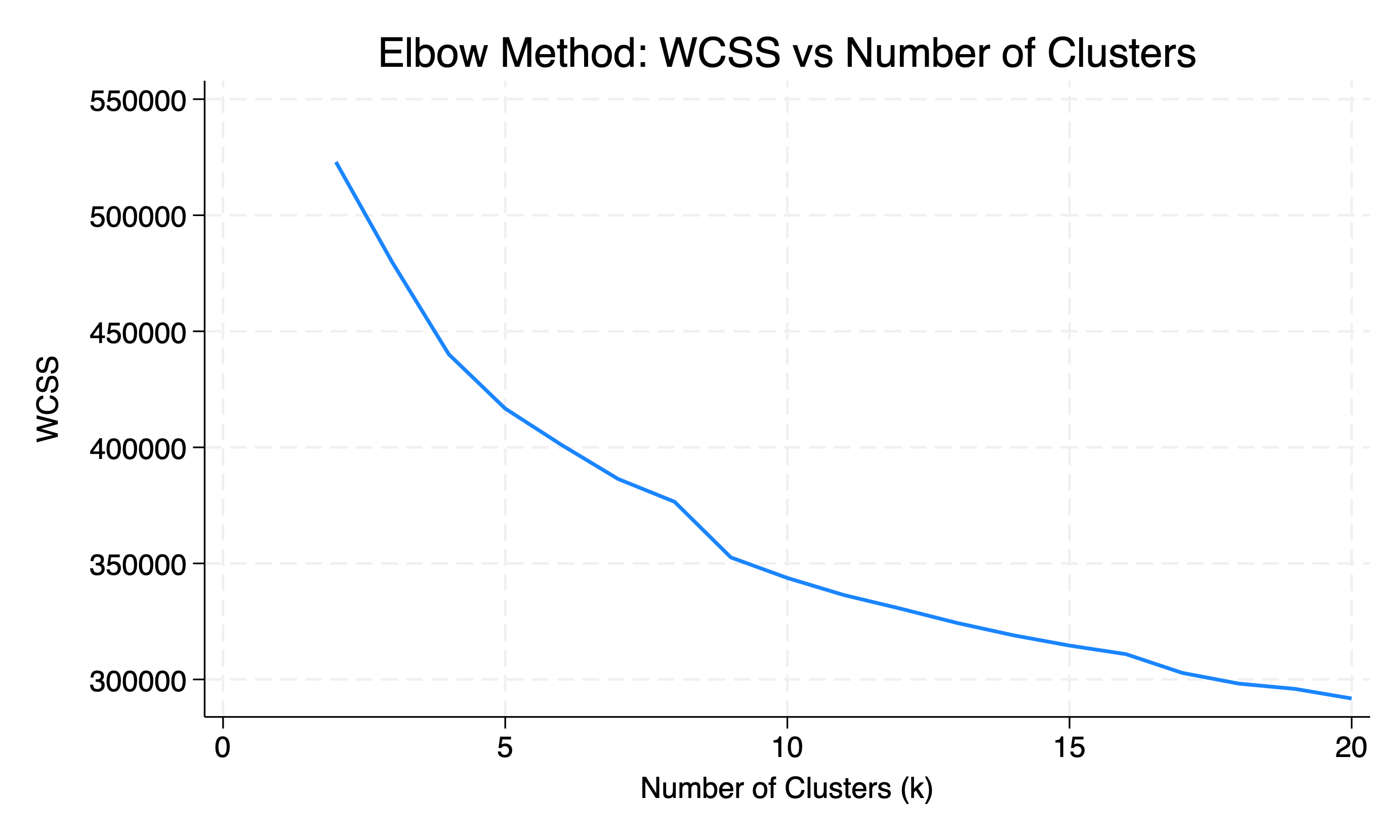

Supplementary Figure 1. Within-clusters sum of squares (WCSS) by number of clusters. All imaging and demographic variables included.

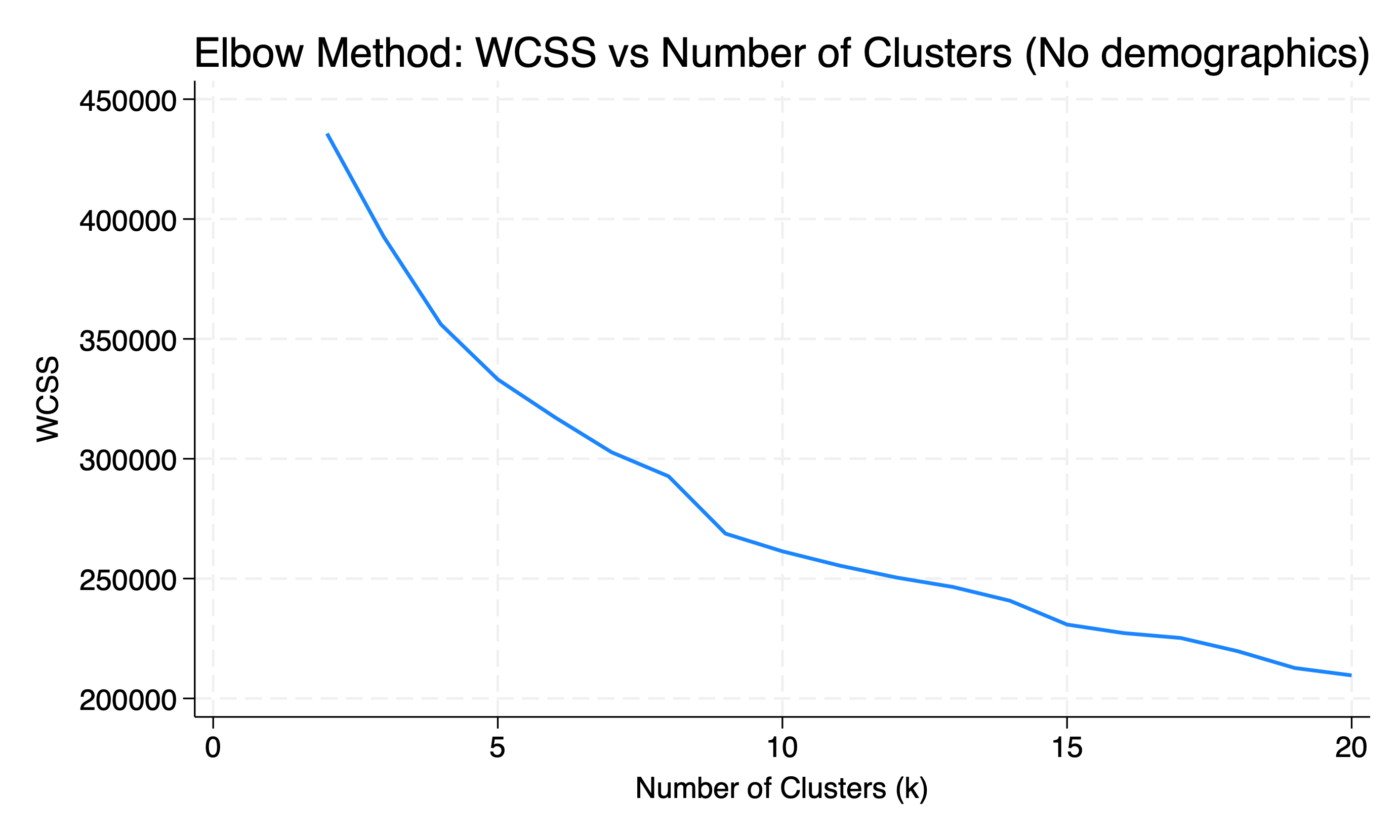

Supplementary Figure 2. Within-clusters sum of squares (WCSS) by number of clusters. All imaging variables included but no demographic variables (i.e., height, weight and age omitted).
