## Supplementary for "Structural phenotypes of osteoarthritis are clinically and genetically distinct: findings from 59,539 UK Biobank participants": Supplementary Results 5.12.25.docx

*Internal validation*

Internal cluster validation revealed acceptable stability across three subsamples. Subsample A showed very high concordance with the primary clustering solution, with a mean overlap of 94%. Subsample C showed lower concordance (56%), largely because bilateral end-stage hip disease in this subsample collapsed into a single cluster due to a higher prevalence of bilateral hip osteoarthritis (12 vs 9%), illustrating that smaller phenotypes are more sensitive to sample composition. Sensitivity analyses that excluded demographic variables also supported nine clusters as the optimal solution (Supplementary Figure 2). Agreement with the main analysis was generally high (mean overlap 86%), except for the body-size phenotype, which showed only 11% overlap because height and weight are central to its definition (Supplementary Table 2).
